# Creation of an Expert Curated Variant List for Clinical Genomic Test Development and Validation: A ClinGen and GeT-RM Collaborative Project

**DOI:** 10.1101/2021.06.09.21258594

**Authors:** Emma Wilcox, Steven M. Harrison, Edward Lockhart, Karl Voelkerding, Ira M. Lubin, ClinGen Expert Panels, Heidi L. Rehm, Lisa Kalman, Birgit Funke

## Abstract

Modern genomic sequencing tests often interrogate large numbers of genes. Identification of appropriate reference materials for development, validation studies, and quality assurance of these tests poses a significant challenge for laboratories. It is difficult to develop and maintain expert knowledge to identify all variants that must be validated to assure analytic and clinical validity. Additionally, it is usually not possible to procure appropriate and characterized genomic DNA reference materials containing the number and scope of variants required. To address these challenges, the Centers for Disease Control and Prevention’s Genetic Testing Reference Material Program (GeT-RM) has partnered with the Clinical Genome Resource (ClinGen) to develop a publicly available list of expert curated, clinically important variants. ClinGen Variant Curation Expert Panels nominated 546 variants found in 84 disease associated genes, including common pathogenic and difficult to detect variants. Variant types nominated included 346 SNVs, 104 deletions, 37 CNVs, 25 duplications, 18 deletion-insertions, 5 inversions, 4 insertions, 2 complex rearrangements, 3 in difficult to sequence regions, and 2 fusions. This expert-curated variant list is a resource that provides a foundation for designing comprehensive validation studies and for creating *in silico* reference materials for clinical genomic test development and validation.

## Introduction

Genetic testing has evolved from interrogating small sets of known, pathogenic variants in one or a few genes using targeted genotyping assays or Sanger sequencing to examining hundreds or thousands of genes at a time using next generation sequencing (NGS). Large gene panels are now the norm in many clinical areas, such as hereditary cancer and cardiomyopathy, and are particularly useful for disorders with locus and allelic heterogeneity. Offerings from molecular testing laboratories range from single disorder to multi-disorder panels, the latter often allowing molecular refinement of the initial clinical diagnosis (National Institutes of Health, Genetic Testing Registry https://www.ncbi.nlm.nih.gov/gtr/ accessed 5/6/2021).

While NGS has advanced many aspects of genetic testing, this technology has made the task of designing and developing analytically and clinically valid tests more complex. This process requires deep clinical and genetic expertise to understand the clinically relevant variant spectrum, and characterization of genomic context as high GC content, highly homologous genes, or repetitive sequences pose challenges for most current NGS technologies .^1, 2^ When examining only one or a few well-studied genes, sufficient expertise to identify clinically important variants can be developed easily, but this aspect of test design and development does not scale to assays that examine hundreds or thousands of genes.^3^ Laboratories often cannot procure reference materials (RMs) that encompass the scope of variants and variant types needed for NGS test development and validation because the supply of available characterized genomic DNA samples from cell lines or patient samples is limited and does not cover the range of genes and variants often included in NGS assays. Finally, traditional patient derived RMs, such as genomic DNA from cell lines, typically contain only 1 or 2 clinically important variants per sample, which can significantly increase the number of samples needed, and thus the complexity and cost of a validation study when large numbers of variants need to be addressed.

Proper analytical validation of genomic tests and large test panels requires a combination of approaches: To ensure optimal confidence in the ability of a given technology to detect variants within a genome or subregion (e.g., exon), large numbers of variants for each clinically relevant type (SNVs, insertions, deletions, SVs) need to be analyzed. In addition, test developers should demonstrate the test’s ability to detect specific clinically important variants particularly when they are technically challenging (Food and Drug Administration https://www.fda.gov/media/99208/download, accessed 3/23/2021; College of American Pathologists, Molecular Pathology 2020 checklist https://www.cap.org/ registration required, accessed 3/23/2021).^4^ Different types of reference materials have been created to meet these needs.

The 1000 Genomes project HapMap samples (1000 Genomes Project
https://www.internationalgenome.org/home; accessed 10/11/2020) were the first generation of characterized genomes that were publicly available. More recently, the National Institute for Standards and Technology (NIST) Genome in a Bottle project has characterized 7 publicly available genomes including NA12878 and two son/mother/father trios of Ashkenazi Jewish and Han Chinese ancestry for use as reference materials (NIST https://www.nist.gov/programs-projects/genome-bottle accessed 5/10/2021).^5, 6^ These samples have been extensively characterized using a number of different technologies and variant calling methods covering over 90% of the genome. NIST has generated publicly available, high confidence benchmarking data sets that have been widely used by the clinical testing community and are now also covering structural variants, and variants in highly homologous regions.^7^ In addition, Illumina produced a set of over 4.7 million phased variants from 17 samples of a large multigenerational pedigree.^8^ These data sets are excellent resources for establishing general analytical performance across various variant types, but do not meet the additional need to validate specific clinically important variants. Other publicly available genomic DNA samples, such as those characterized by the Center for Disease Control and Prevention’s Genetic Testing Reference Material Program (GeT-RM https://www.cdc.gov/labquality/get-rm/index.html accessed 10/11/2020) and samples available from repositories such as Coriell Institute for Medical Research (Coriell https://www.coriell.org/ accessed 5/10/2021) or the American Type Culture Collection (ATCC) (ATCC https://www.atcc.org/ accessed 5/10/2021) have identified variants in some, but not all genes likely to be included in NGS tests. Although useful, none of these genomic DNA samples, individually or as a group, are sufficient to represent the wide range of variants needed for a thorough and comprehensive validation of an NGS assay.

As NGS tests have grown to include hundreds to thousands of genes, it has become technically and economically impractical to rely exclusively on DNA samples for assay development and validation studies to assess assay performance. Procuring sufficient reference materials containing specific variants has always been challenging and can be virtually impossible for large gene panels, particularly those that span multiple disorders. In addition, the commoditization of genomic testing and the availability of “off-the-shelf” gene panels have facilitated implementation of genomic tests. -Well established laboratories with longstanding clinical expertise often have archived DNA which can be used as reference materials for test development. In contrast, laboratories that do not already have in-depth expertise in the genomic profile of the disorders included in the panels are faced with a paucity of organized resources and guidance as to which clinically important variants require specific attention during test development. The FDA and professional organizations including the College of American Pathologists have begun to acknowledge these challenges and have responded with guidance that permits use of *in silico* reference materials to supplement clinical test validation and operation (Food and Drug Administration https://www.fda.gov/media/99208/download, accessed 3/23/2021; College of American Pathologists, Molecular Pathology 2020 checklist https://www.cap.org/ registration required, accessed 3/23/2021).

In 2019, the Clinical Laboratory Improvement Advisory Committee (CLIAC) issued a recommendation to the Department of Health and Human Services that the GeT-RM program should develop electronic reference materials for NGS testing (https://www.cdc.gov/cliac/docs/summary/cliac0419_summary.pdfaccessed5/10/2021). As a first step towards this goal, GeT-RM has partnered with the Clinical Genome Resource (ClinGen, https://www.clinicalgenome.org/ accessed 2/27/2021) to develop lists of expert curated, clinically important variants. This publication describes these variant lists, which can be used to inform test validation studies and will serve as a foundation for creating multi-variant electronic reference materials by *in silico* mutagenesis of laboratory generated BAM or FASTQ files.^9-11^

## Methods

### Variant nomination process

Members of 36 expert panels, including 35 of ClinGen’s Variant Curation Expert Panels (VCEPs), and experts from the CFTR2 Project were contacted in March 2020 and asked to nominate clinically important variants in the genes covered by the expert panels. FDA has recognized ClinGen’s curation process and its resulting variant classifications as a regulatory-grade variant database (ClinGen https://clinicalgenome.org/docs/fda-recognizes-clingen-assertions-in-clinvar-frequently-asked-questions/ accessed 5/10/2021; Food and Drug Administration https://www.fda.gov/news-events/press-announcements/fda-takes-new-action-advance-development-reliable-and-beneficial-genetic-tests-can-improve-patient accessed 5/10/2021). These VCEP curated, FDA recognized variants are available via NCBI’s ClinVar database and ClinGen’s Evidence Repository (https://erepo.clinicalgenome.org accessed 5/10/2021).

Each expert panel was asked to nominate a minimum of 10 variants per disease area, including pathogenic variants that are the largest contributors to a disease as well as variants that may represent potential analytic challenges to laboratories. Panels submitted their variant nominations on a standardized template from April-June 2020. For each variant, the panels noted if the basis for inclusion was: a) Major contributor to disease, b) Analytic detection difficult, c) Filtration impact (may be inadvertently filtered out due to high allele frequency), or d) Other (with explanation provided in a comment box). The panels were instructed that any reason for difficult detection was appropriate including: variant type, homology issues or a high allele frequency for a lower penetrant variant that might make the variant excluded by standard allele frequency filters. The ClinGen Allele Registry was used to standardize nomenclature for all nominated variants and ClinVar Variation IDs and associated disorders were added where available. A full list of the expert panels that contributed variants can be found in Supplemental Table 1 and the full list of nominated variants can be found in Supplemental Table 2. Not all of the ClinGen VCEPs that participated in this project have completed all four steps of the VCEP development process required for final approval by ClinGen^12^ (ClinGen https://clinicalgenome.org/docs/guidelines-for-applying-for-variant-or-gene-curation-expert-panel-status/ accessed 3/23/2021). Of the variants submitted by the panels, 38% are from expert panels with final ClinGen approval to submit to ClinVar as ‘reviewed by expert panel’, whereas 62% are from ClinGen VCEPs that were still developing disease specifications for variant classification and were not yet fully ClinGen-approved at the time of submission of variants to the project. VCEP status at the time of manuscript submission is shown in column B of Supplemental table 2. Current status can be found on the ClinGen website (https://clinicalgenome.org/working-groups/clinical-domain/ accessed 3/23/2021).

### Limitations of variant selection process

Several limitations of the variant selection process should be noted. There were no explicit requirements given to the expert panels in terms of the definitions of categories a, b, and c and therefore each may have used slightly different definitions to select variants to nominate for this project. In addition, some expert panels may have used laboratory data that is not in the public domain to identify nominated variants and this information was not requested during the submission process. Furthermore, some expert panels represent genes and diseases that are very rare and therefore variants may not, in general, be highly prevalent. These are not considered serious limitations as these are “first-generation” variant lists, which provide an entirely new resource. It is expected that this effort will continue in years to come to broaden and deepen this resource.

## Results

Nominations of clinically important pathogenic or difficult to detect variants were received from 24 expert panels out of 36 contacted (Supplemental Table 1). Overall, 546 unique variants in 84 genes were nominated (Supplemental Table 2) including 22 of the 59 genes recommended by ACMG for reporting of incidental or secondary findings, which are included as part of virtually all proactive genomic health tests.^12^ The nominated variants are causative for a wide range of diseases many of which are commonly tested by NGS including heritable cancers, inborn errors of metabolism, cardiomyopathy, diabetes, and immune disorders (Table 1). A number of different variant types were nominated, including 346 SNVs, 104 deletions, 37 CNVs, 25 duplications, 18 deletion-insertions, 5 inversions, 4 insertions, 2 complex rearrangements, 2 fusions, and 3 variants in regions defined as “difficult region to cover and map variants”. The variants were nominated for a variety of reasons: 355 are major contributors to disease, 111 are analytically difficult to detect, 28 may have filtration impacts (be inadvertently filtered out due to high allele frequency), 6 had multiple reasons for inclusion, and 46 were listed as “other” or no explanation was provided (Supplemental Table 2).

**Table 1.**
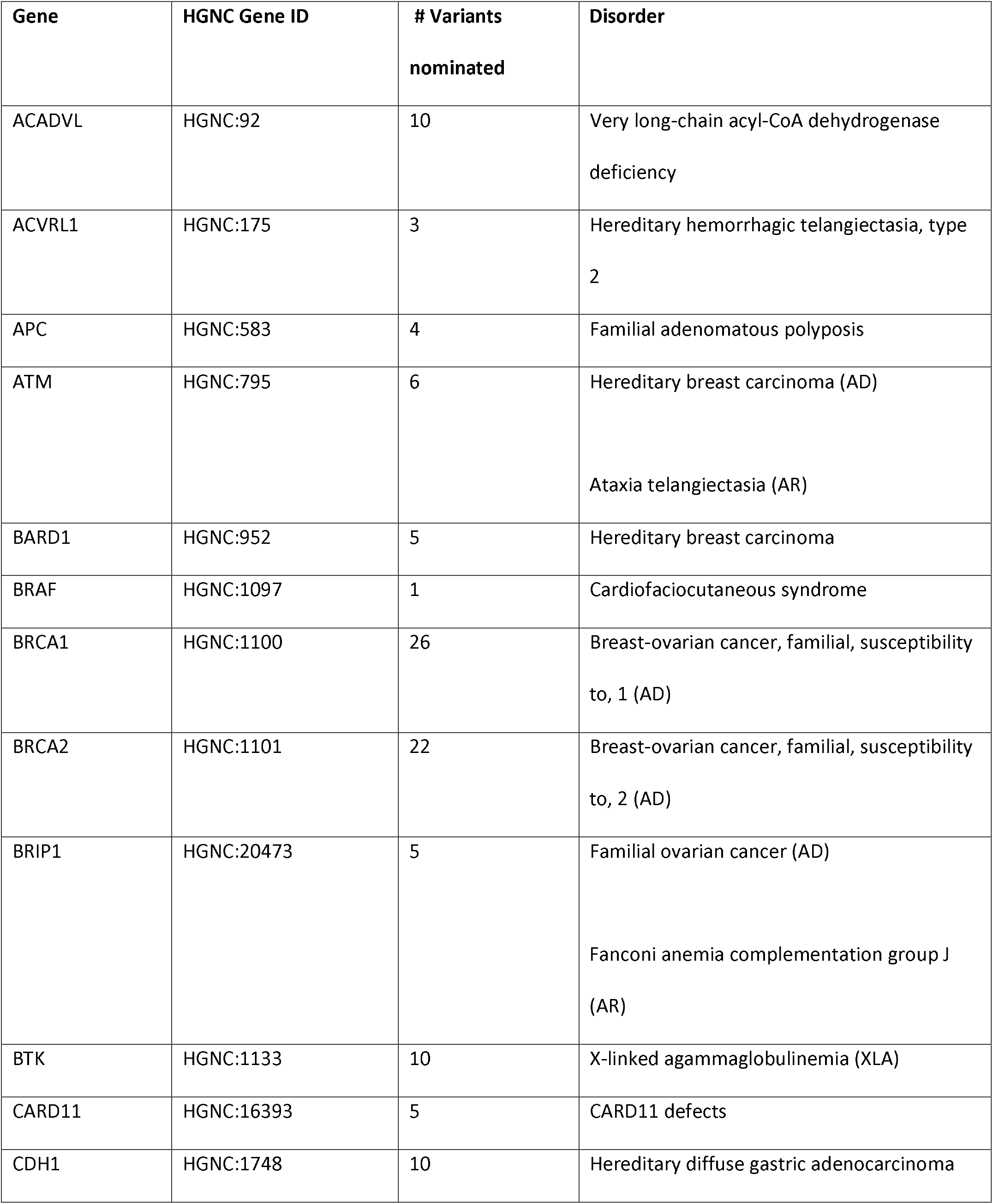
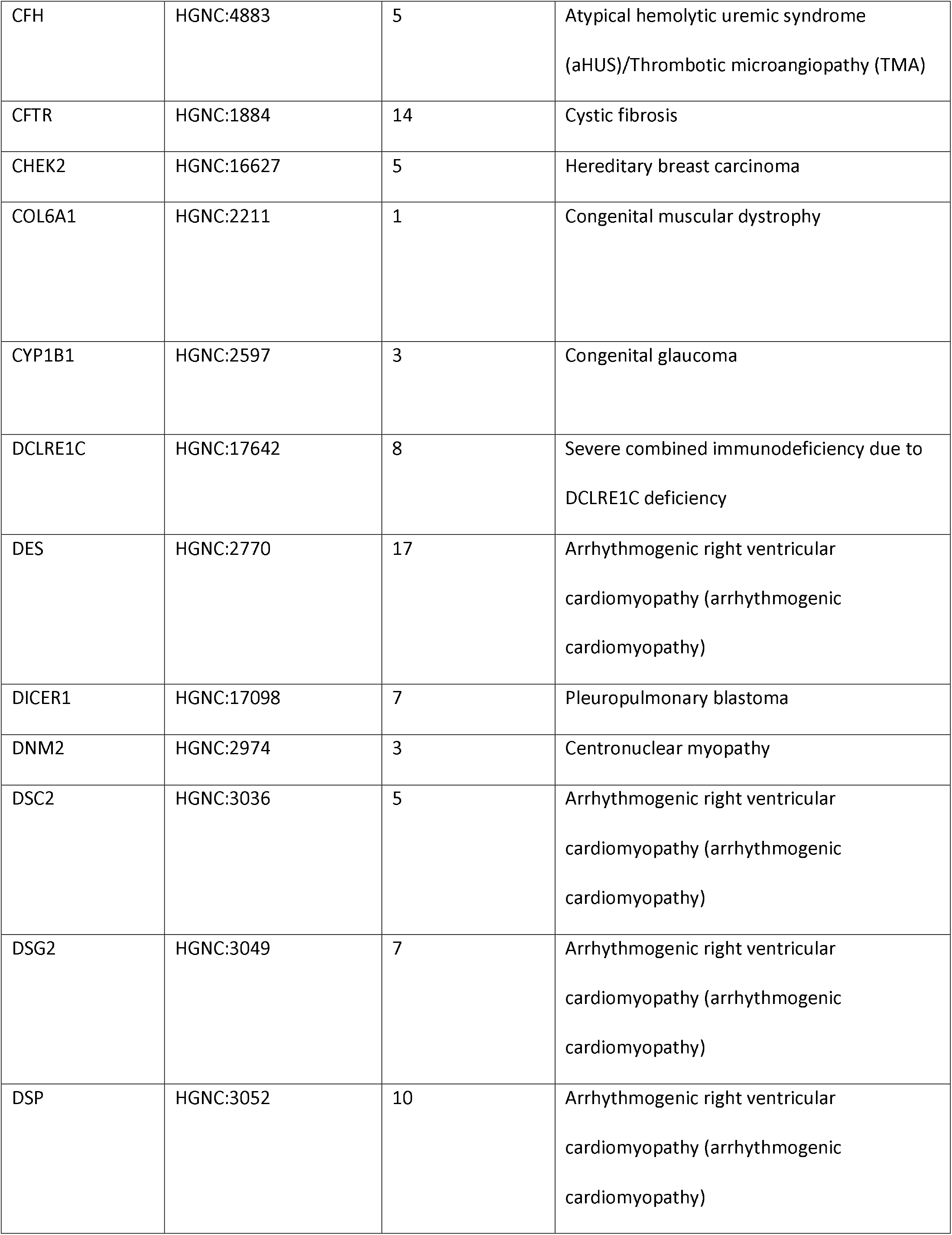
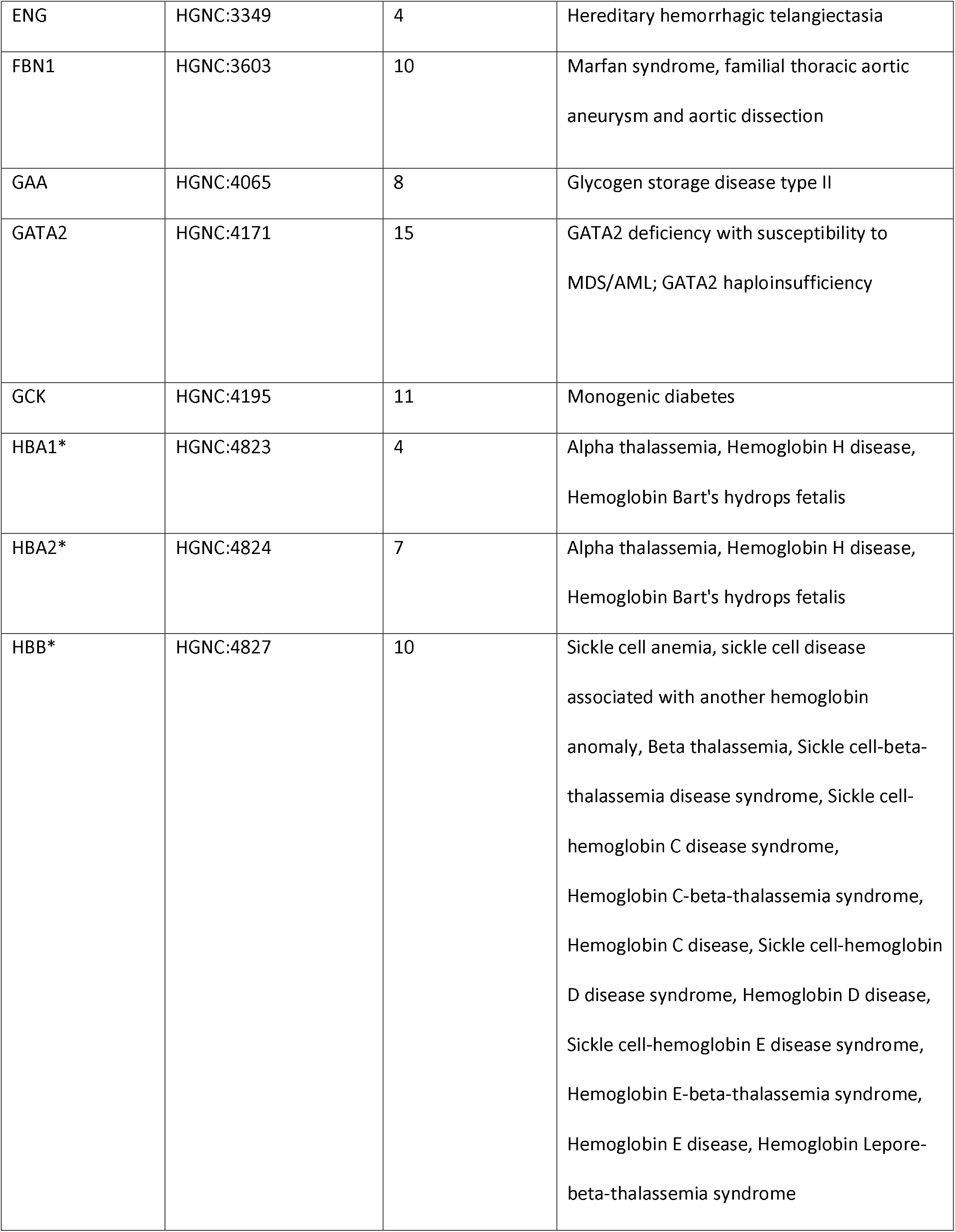
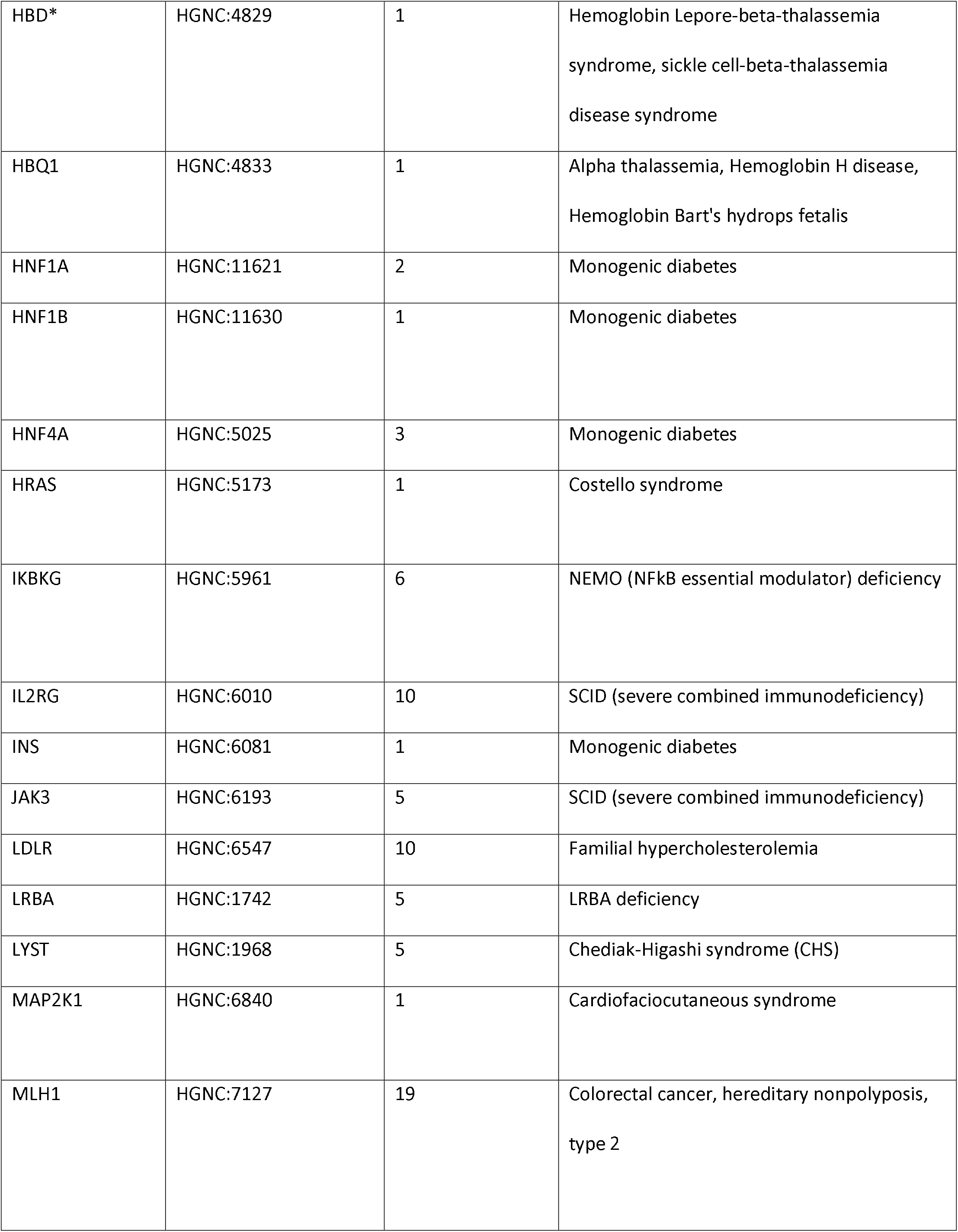
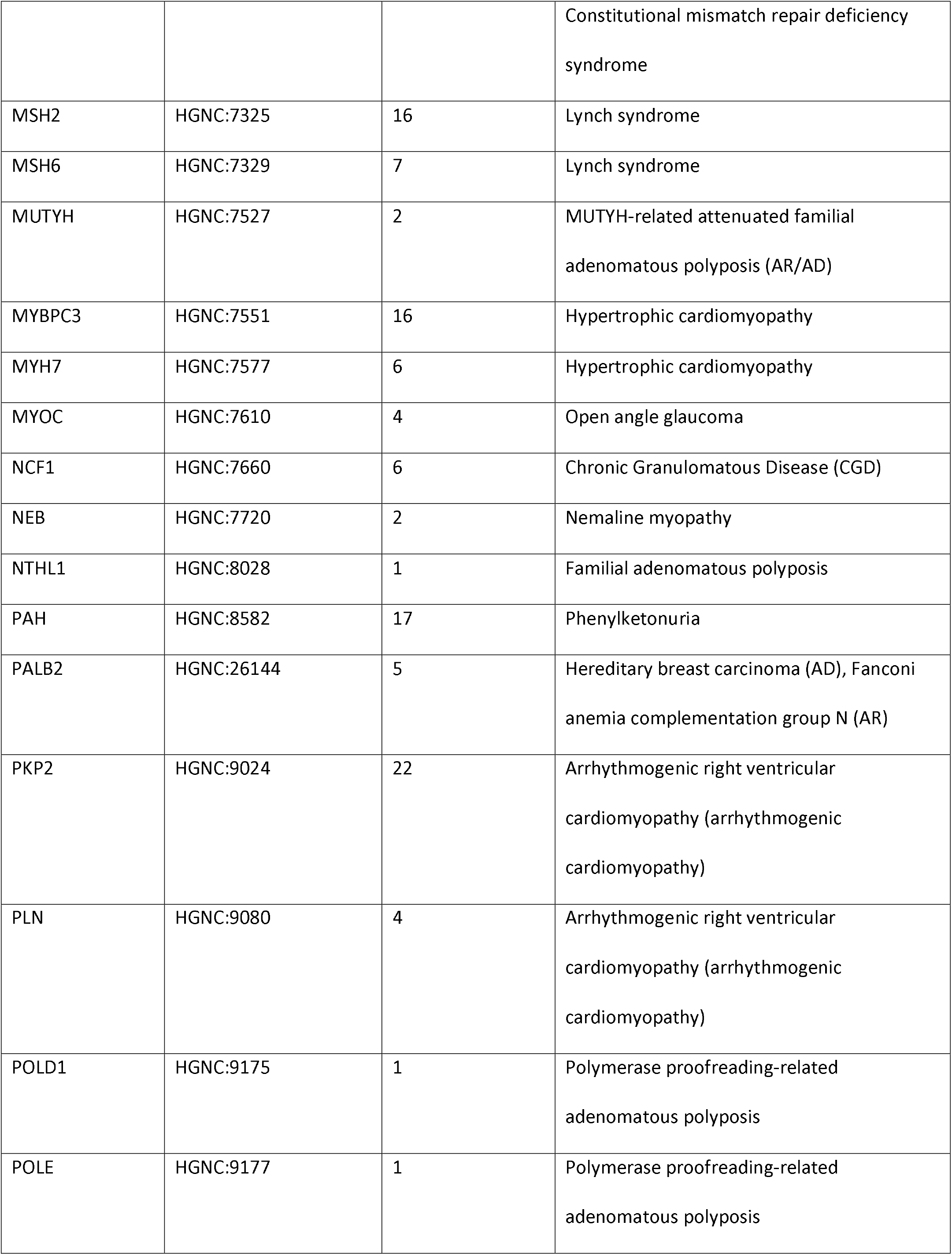
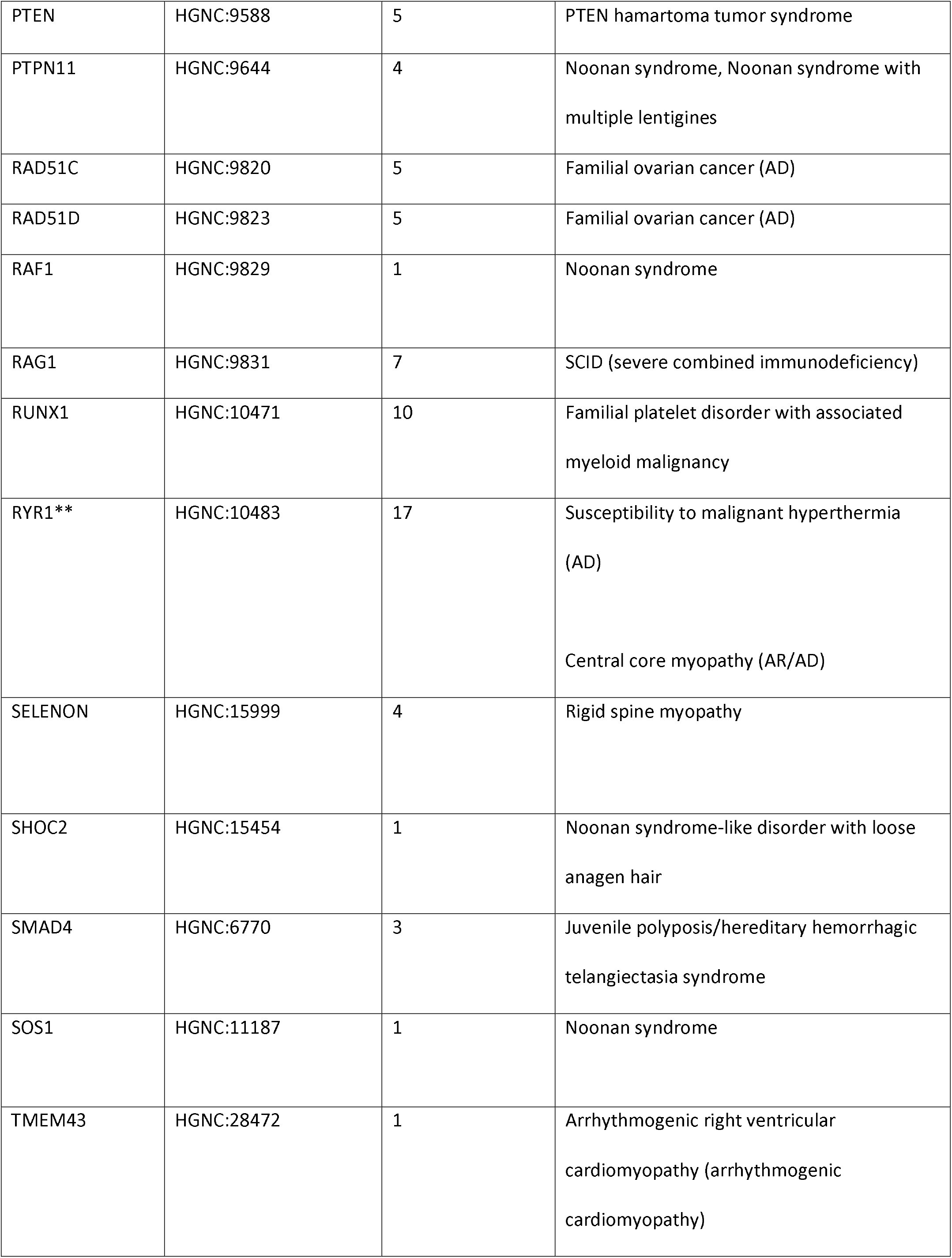
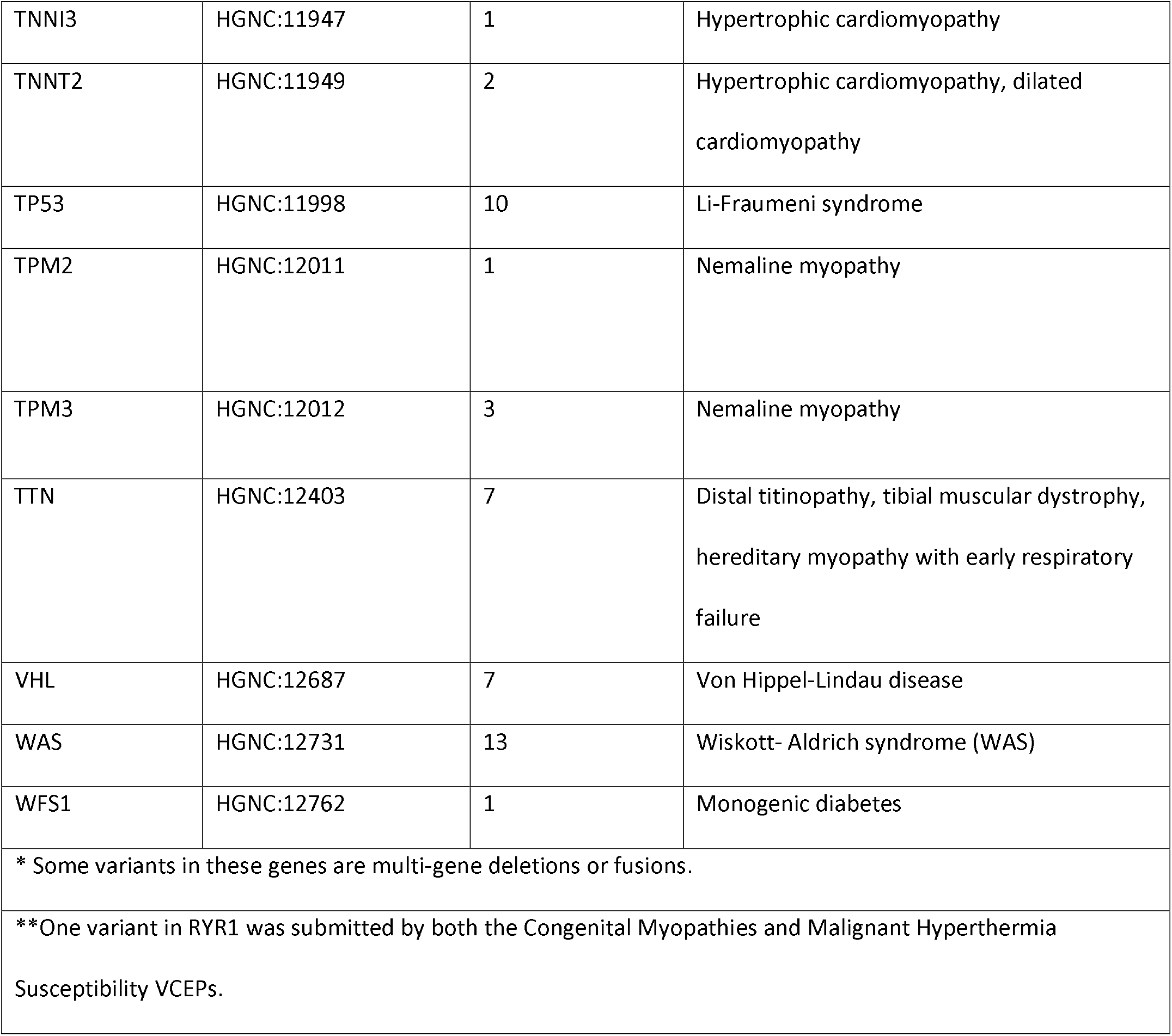
Summary of the numbers of variants per gene nominated

## Discussion

Clinical laboratories in the United States are required by regulation and guided by professional or best practice standards to use characterized reference materials for test development, validation and verification studies, quality control and proficiency testing^13-16^ (American College of Medical Genetics https://www.acmg.net/PDFLibrary/Standards-Guidelines-Clinical-Molecular-Genetics.pdf, accessed 4/9/2020, Washington State Legislature, http://app.leg.wa.gov/WAC/default.aspx?cite=246-338-090, accessed 4/9/2020, College of American Pathologists https://www.cap.org/, accessed 4/9/2020 (registration required), New York State Clinical Laboratory Evaluation Program, http://www.wadsworth.org/clep, accessed 4/9/2020).

Certain aspects of test development and validation, such as DNA extraction, library preparation, and sequence generation will always require the use of genomic DNA from cell lines or patient samples. However, given the scope of many NGS tests, it is no longer feasible to rely solely upon genomic DNA samples containing only one or a few variants of interest to validate an assay that tests an entire exome or genome or many genes within a large NGS panel, and novel types of reference materials are needed to enable assessment of a test’s analytical validity and establish adequate clinical validity.

The Association for Molecular Pathology and the College of American Pathologists allow the use of *in silico* reference materials to supplement genomic DNA for the validation of bioinformatic pipelines (College of American Pathologists, Molecular Pathology 2020 checklist https://www.cap.org/ registration required, accessed 3/23/2021).^4, 17-19^ Specialized software^9, 10, 20^ has been developed to edit sequence variants into BAM or FASTQ files generated by the assay being validated to create customized reference materials. The software is publicly available, and several companies offer this service to laboratories. The GeT-RM program is planning to use these curated variants and file editing software to pilot development and use of *in silico* reference materials in clinical laboratories.

The number of variants that can be added is potentially unlimited, but care must be taken to avoid adding too many variants in *cis* to the same sequence reads because it may affect alignment to the reference sequence. The types of variants that can be introduced remains limited to SNVs and smaller insertions and deletions at different allele fractions, although the ability to introduce CNVs is currently being developed. This approach has been used successfully by the College of American Pathologists for its NGS proficiency testing challenges.^9, 10^

The variant list developed by the effort presented here is available on the GeT-RM and ClinGen websites and can serve as a knowledge resource to help laboratories identify important pathogenic and difficult to detect variants in genes that are included in their assays, as well as a foundation for generating customized *in silico* reference materials. GeT-RM and ClinGen will continue to collaborate to add to the current variant list and invite input from the genetics community about this list and the processes used to generate it.

## Data Availability

All data is included in the figures, tables and supplement included in the manuscript.

## Acknowledgements

ClinGen is primarily funded by the National Human Genome Research Institute (NHGRI), through the following three grants: U41HG006834, U41HG009649, U41HG009650. ClinGen also receives support for content curation from the Eunice Kennedy Shriver National Institute of Child Health and Human Development (NICHD), through the following three grants: U24HD093483, U24HD093486, U24HD093487.

---

Suppl Table 1. List of participating expert panels and names of participants

Suppl Table 2. List of variants nominated

